# Simulation of undiagnosed patients with novel genetic conditions

**DOI:** 10.1101/2022.08.22.22279073

**Authors:** Emily Alsentzer, Samuel G. Finlayson, Michelle M. Li, Undiagnosed Diseases Network, Shilpa N. Kobren, Isaac S. Kohane

## Abstract

Rare Mendelian disorders pose a major diagnostic challenge and collectively affect 300-400 million patients worldwide. Many automated tools aim to uncover causal genes in patients with suspected genetic disorders, but evaluation of these tools is limited due to the lack of comprehensive benchmark datasets that include previously unpublished conditions. Here, we present a computational pipeline that simulates realistic clinical datasets to address this deficit. Our framework jointly simulates complex phenotypes and challenging candidate genes and produces patients with novel genetic conditions. We demonstrate the similarity of our simulated patients to real patients from the Undiagnosed Diseases Network and evaluate common gene prioritization methods on the simulated cohort. These prioritization methods recover known gene-disease associations but perform poorly on diagnosing patients with novel genetic disorders. Our publicly-available dataset and codebase can be utilized by medical genetics researchers to evaluate, compare, and improve tools that aid in the diagnostic process.

## Introduction

Rare congenital disorders are estimated to affect nearly 1 in 17 people worldwide [1], yet the genetic underpinnings of these conditions—knowledge of which could improve support and treatment for scores of patients—remain elusive for 70% of individuals seeking a diagnosis and for half of suspected Mendelian conditions in general [2, 3]. This diagnostic deficit results in a substantial cumulative loss of quality-adjusted life years and a disproportionate burden on healthcare systems overall [4–6]. Organizations such as the Undiagnosed Disease Network (UDN) in the United States have been established to facilitate the diagnosis of such patients, which has resulted in both successful diagnoses for many patients as well as the discovery of new diseases [7, 8].

The diagnostic workup of patients with suspected Mendelian disorders increasingly includes genomic sequencing. Whole genome or exome sequencing typically identifies thousands of genetic variants which must be analyzed to identify the subset of causal variant(s) yielding the patient’s syndrome (Figure 1a). This process is challenging and error prone; for example, patients may have variants that do not ultimately cause their presenting syndrome, yet fall into genes that are plausibly associated with one or more of their phenotypes. Further challenges arise in situations where patients present with a novel set of symptoms that do not match any known disorder, or when their disease-causing variants occur in genes not previously associated with any disease (Figure 1b). In the first phase of the UDN for instance, 23% of eventual patient diagnoses were due to novel syndromes [4].

**Figure 1:**
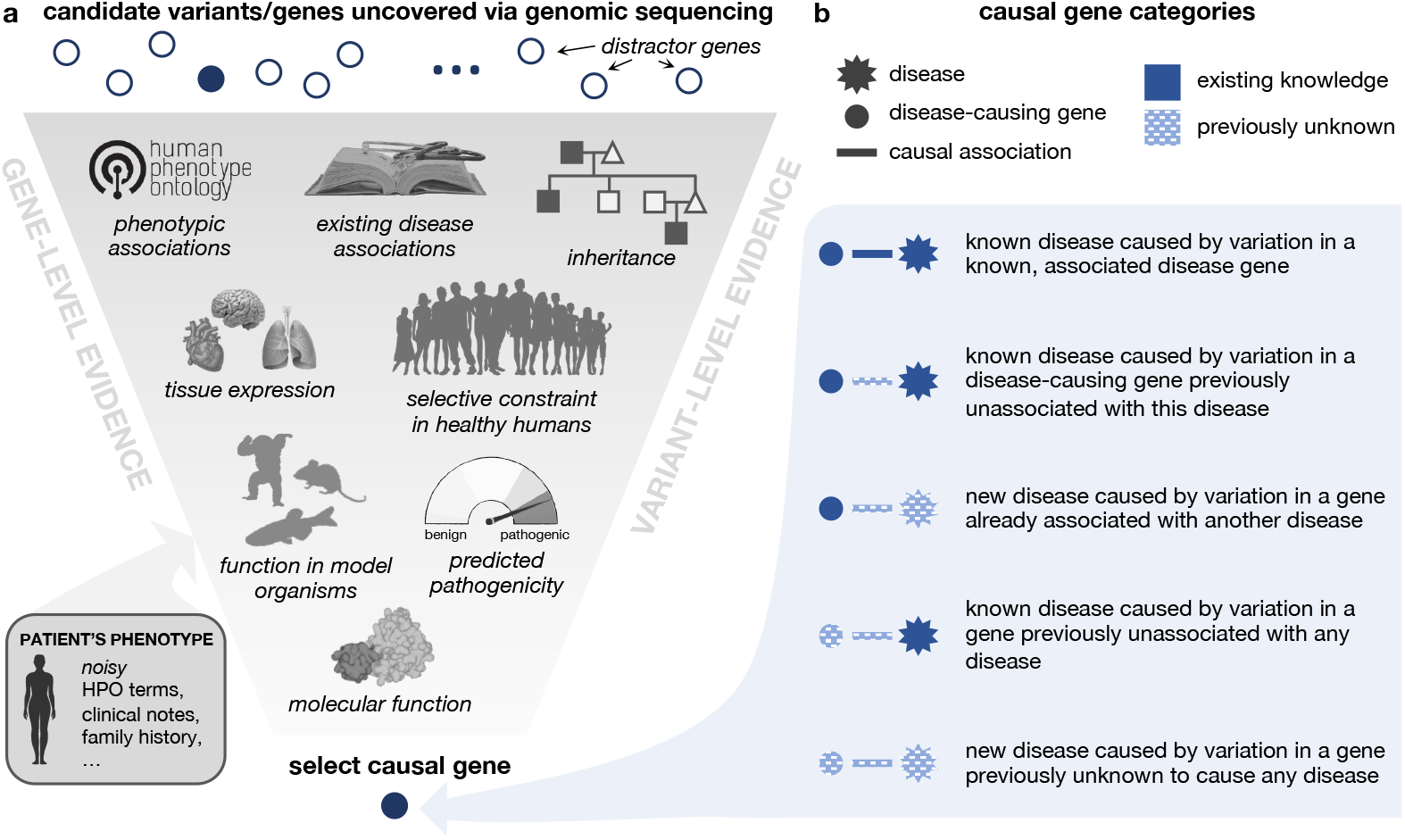
Identification and categorization of causal disease genes. **a**. Genomic variation uncovered in an affected patient through DNA sequencing is investigated using variant-level and gene-level evidence in order to identify the impacted gene that is most likely responsible for causing the patient’s symptoms. Here we depict a subset of relevant information that a care team may use to make this assessment. **b**. The causal gene responsible for a patient’s disorder can be categorized based on the extent of medical knowledge that exists about the gene and its associated disorder. Intuitively, diagnosing patients where less is known about their causal gene and disease (bottom category) is a more challenging task than diagnosing patients where more is known about causal gene and disease (top category).

To accelerate the diagnostic process, a plethora of computational tools are used by clinical teams to automatically analyze patients’ genetic and phenotypic data to prioritize causal variants [9–12]. Unfortunately, a comprehensive evaluation of these tools’ performance is hindered by the lack of a public benchmark database of rare disease patients of sufficient size to cover the full breadth of genomic diseases. While efforts such as the Deciphering Developmental Disorders project provide useful benchmarks for specific populations of rare disease patients, they are limited in diagnostic scope and require an extensive DUA for full access [13]. In lieu of real patient data, simulated patient data offers several clear advantages: the simulation approach can be scaled to an arbitrary number of patients and disorders, data are inherently private, and the transparency of the simulation process can be leveraged to expose specific failure modes of different methods. However, simulated patient data is only useful insofar as it reflects the challenges of real-world diagnosis. This requires a faithful simulation of the complex relationship between candidate genes and the patient’s phenotypes as well as the notion of disease novelty, as described above. Existing approaches for simulating rare disease patients unrealistically model the patient’s genotype and phenotype disjointly by inserting disease-causing alleles into otherwise healthy exomes and separately simulating patient phenotypes as a set of precise, imprecise, and noisy phenotypes [14–16]. Although healthy individuals may randomly harbor variants in disease-causing genes, these genes would be weak candidates as the patients’ symptoms would not cause a physician to hone in on those genomic regions. In addition, with few exceptions, most studies analyzing tools for rare disease diagnosis do not assess the ability of tools to identify novel syndromes or variants [15, 17, 18]. Given the importance and prevalence of automated prioritization tools in the diagnostic process, enabling meaningful comparisons and improvement of these tools via benchmarks that capture the notion of novelty and realistic phenotypes will be essential.

Here, we present a computational pipeline to simulate undiagnosed patients that can be used to evaluate gene prioritization tools. Each simulated patient is represented by sets of candidate disease-causing genes and standardized phenotype terms. To model novel genetic conditions in our simulated patients, we first curate a knowledge graph (KG) of known gene–disease and gene–phenotype annotations that is time-stamped to 2015. This enables us to define post-2015 medical genetics discoveries as novel with respect to our KG. We additionally provide a taxonomy of categories of “distractor” genes that do not cause the patient’s presenting syndrome yet would be considered plausible candidates during the clinical process. We then introduce a simulation framework that jointly samples genes and phenotypes according to these categories to simulate nontrivial and realistic patients and show that our simulated patients closely resemble real-world patients profiled in the UDN. Finally, we reimplement existing gene prioritization algorithms and assess their performance in identifying etiological genes in our simulated patient set, revealing specific settings in which established tools excel or fall short. Overall, the approach to patient simulation we present here is, to the best of our knowledge, the first to incorporate realistic, non-trivial candidate distractor genes and phenotype annotations as well as the notion of novel genetic disorders. We provide our framework and simulated patients as a public resource to advance the development of new and improved tools for medical genetics.

## Results

We design and implement a pipeline for simulating patients with difficult-to-diagnose Mendelian disorders (Figure 2a). Each simulated patient is represented by an age range, a set of positive symptoms (phenotypes) that they exhibit, a set of negative phenotypes that they do not exhibit, and a set of candidate genes that may be causing their disease. There are three components of our simulation framework. First, each patient is initialized with a genetic disorder profiled in the comprehensive and well-maintained rare genetic disease database Orphanet [19]. Second, the imprecision in real-world diagnostic evaluations is modeled via *phenotype dropout* to mimic patients’ partially observed symptoms, *phenotype corruption* which replaces specific symptoms with more general phenotype terms, and *phenotype noise* which adds unrelated symptoms and comorbidities proportionally to their prevalence in age-matched patients from a medical insurance claims database. Finally, we develop a framework to generate strong, yet ultimately noncausal, candidate genes inspired by the typical rare disease diagnostic process (Figure 2b). These challenging distractor genes and some associated phenotype terms are added according to each of six distractor gene modules (See Methods for further details).

**Figure 2:**
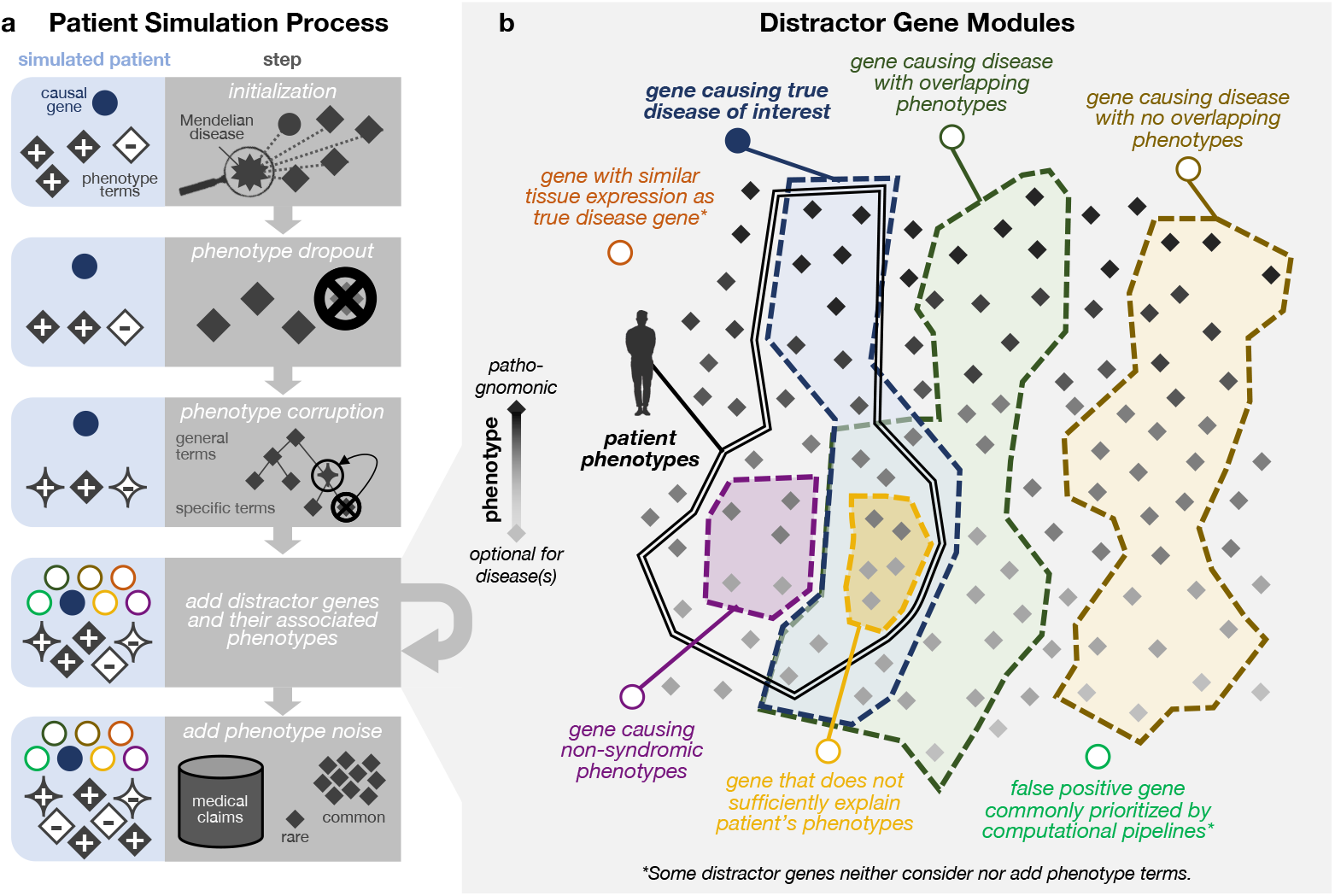
Simulation process generates patients with multiple phenotype terms and candidate genes. **a**. Patients are first assigned a true disease and initialized with a gene known to cause that disease (blue circle) as well as with positive and negative phenotypes associated with that disease (gray diamonds). Phenotype terms are then randomly removed through phenotype dropout, randomly altered to be less specific according to their position in an ontology relating phenotype terms, and augmented with terms randomly selected by prevalence in a medical claims database. Finally, strong distractor candidate genes and relevant additional phenotypes are generated based on six distractor gene modules. **b**. The six distractor gene modules are inspired by genes that are frequently considered in current clinical genomic workflows and are designed to generate highly plausible, yet ultimately non-causal, genes for each patient. Four of the distractor gene modules are defined by the overlap—or lack thereof —between the phenotypes associated with the distractor gene and the phenotypes associated with the patient’s causal gene. The remaining two modules are defined by their similar tissue expression as the true disease gene or solely by their frequent erroneous prioritization in computational pipelines.

### Simulated patients mimic real-world patients

We leverage our computational pipeline to simulate 42,680 realistic rare disease patients representing a total of 2,134 unique Mendelian disorders and 2,401 unique causal genes. Each simulated patient is characterized by 18.39 positive phenotypes (*sd* = 7.7), 13.5 negative phenotypes (*sd* = 8.5), and 14 candidate genes (*sd* = 3.5) on average.

To assess whether our simulated patients are systematically distinguishable from real-world patients, we assemble a cohort of 121 real-world patients from the Undiagnosed Diseases Network (UDN) who were diagnosed with a disease in Orphanet annotated with genes and phenotypes and then select 2,420 simulated patients with matching diseases. There are 92 unique diseases represented in the real and disease-matched simulated patient cohorts. Real and simulated patients have similar numbers of candidate genes (13.13 vs 13.94 on average; Figure 3a) and positive phenotype terms (24.08 vs 21.57 on average; Figure 3b). Real-world patients are also more similar to their simulated counterparts than to other real-world patients. When we apply dimensionality reduction on the positive phenotype terms of patients, real-world patients cluster with and are visually indistinguishable from simulated patients within each disease category (Figure 3c), suggesting that there are not consistent differences in phenotype term usage between real and simulated patients. Moreover, for each real-world patient, the ten phenotypically-closest simulated patients with the same disease are closer than the ten phenotypically-closest real-world patients with different diseases (average Jaccard Similarity of 0.952 vs 0.930; p-value=7.4e-81, Wilcoxon one-sided test). We also employ a nearest neighbor analysis using Jaccard Similarity as our distance metric to evaluate whether the simulated patients’ phenotype terms are sufficiently reflective of and specific to their assigned diseases. We find that simulated patients with similar sets of phenotype terms to real patients are more likely to have the same disease as those real patients than randomly selected simulated patients (Figure 3d).

**Figure 3:**
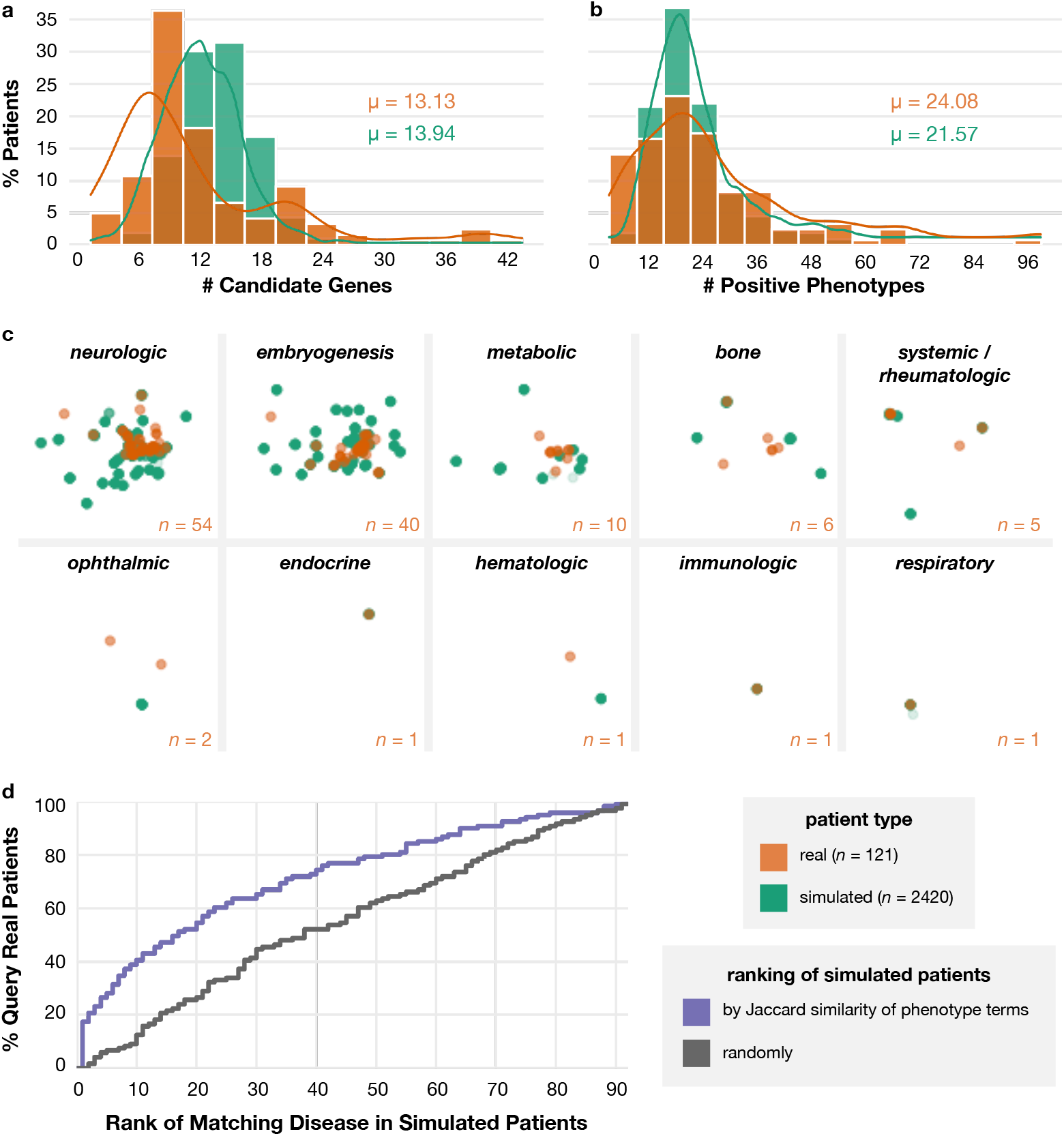
Simulated patients mimic real-world patients. Diagnosed, real-world patients from the Undiagnosed Diseases Network (orange) and a disease-matched cohort of simulated patients (teal) have similar numbers of **a** candidate genes per patient (average of 13.13 vs. 13.94) and **b** positive phenotype terms per patient (average of 24.08 vs. 21.57). **c**. Real patients (orange) and simulated patients (teal) are indistinguishable based on their annotated positive phenotype terms within each Orphanet disease category, as visualized using non-linear dimension reduction via a Uniform Manifold Approximation and Projection (UMAP) plot. The horizontal and vertical axes are uniform across all plots. The number of real patients within each disease category is listed in the corner of each plot; there are 20 simulated patients for each real patient. **d**. For each real-world patient, all simulated patients in the disease-matched cohort are ranked randomly (black) and by the Jaccard similarity of their phenotype terms to the query real-world patient (purple). The Cumulative Distribution Function (ECDF) plot shows that the basic Jaccard similarity metric is able to retrieve simulated patients with the same disease as the query real patient more accurately than if the simulated patients were retrieved randomly.

### Pipeline simulates patients with novel and diverse genetic conditions

A primary challenge in diagnosing real-world patients, and one that should be reflected in relevant simulated patients, is when their causal gene-disease relationships have never previously been documented (Figure 1b). However, simulating patients with “novel” genetic conditions is conceptually nontrivial, as simulated disease associations must be drawn from some existing knowledge graph. To overcome this issue, we consider the gene-phenotype-disease associations annotated in a knowledge graph timestamped to 2015 to be “existing knowledge” and discoveries made post-2015 to be “novel” (see Section 6). This enables us to categorize simulated patients according to the novelty of their gene-disease relationships with respect to this timestamped knowledge graph (Table 1). Although only 2% and 1% of the total simulated patients respectively correspond to previously known and previously unknown diseases caused by genes never before associated with any disease, the total number of simulated patients in these two categories are 14**x** and 190**x** higher respectively than in our real-world dataset. Moreover, whereas only 231 unique disease genes have been identified as causal in our phenotypically-diverse, real-world UDN dataset, our simulated patients’ 2,100+ unique diseases are caused by 2,401 unique disease genes. Overall, these results demonstrate that our pipeline can simulate substantially higher numbers of patients—that are diverse with respect to disease and the degree of novelty of their causal gene-disease relationships—than compared to a national dataset of real-world patient data.

**Table 1:** Counts of simulated and real-world UDN patients in each causal gene–disease category. UDN patients with multiple causal genes may appear in several categories.

| Causal Gene Category | # Simulated Patients | # Real-world UDN Patients |
| --- | --- | --- |
| Known disease caused by an associated causal gene | 36,226 (85%) | 149 (58%) |
| Known disease caused by a disease-causing gene previously unassociated with their disease | 4,339 (10%) | 15 (6%) |
| Known disease caused by a gene never before associated with any disease | 835 (2%) | 5 (2%) |
| Novel disease caused by a disease-causing gene | 947 (2%) | 60 (23%) |
| Novel disease caused by a gene never before associated with any disease | 333 (1%) | 29 (11%) |

### Performance of gene prioritization algorithms on real and simulated patients

The size and diversity of our real and simulated patient datasets enable us to evaluate how well existing algorithms are able to prioritize causal genes in patients with different degrees of preexisting knowledge of their gene–disease relationships. We run four commonly-used gene prioritization algorithms on patients in each of the causal gene–disease association categories outlined in Figure 1b, ensuring that each algorithm only had access to the existing knowledge found in the timestamped 2015 knowledge graph. Each algorithm inputs the patient’s positive phenotype terms and the patient’s individualized set of candidate genes and produces a ranking of the candidate genes according to how likely they are to cause the patient’s phenotypes. Briefly, Phrank uses semantic similarity of phenotype terms in the Human Phenotype Ontology (HPO) and prevalence of gene associations across these phenotypes to match patient symptoms to genes or diseases [10]. The Patient–Gene version of Phrank directly compares a patient’s phenotype terms and the phenotype terms associated with the candidate gene. The Patient-Disease version considers all diseases associated with a candidate gene and compares the patients’ phenotypes and those diseases’ phenotypes, assigning the candidate gene the highest similarity score across all of its associated diseases. Phenomizer uses semantic similarity of phenotype terms and prevalence of phenotype associations across all known diseases to match patient symptoms to diseases [12]. Candidate genes are assigned the highest score across their associated diseases. Phenolyzer uses semantic similarity to match patient symptoms to diseases and scores genes directly associated with the diseases as well as additional genes connected via a gene-gene network [11].

### Real-world and simulated patients are equally difficult to diagnose

We first assess whether gene prioritization performance was similar between the simulated patient cohort and the real-world patient cohort. Across both patient sets, correctly ranking the causal gene becomes more difficult as the amount of information about the causal gene-disease relationship in the knowledge graph decreases (Figure 4). We find that the performance between simulated and real-world patients was similar for all four algorithms across all but the easiest gene-disease association categories. Overall, these results indicate that the simulated patient cohort can serve as a reasonable proxy for real-world patients, particularly when evaluating a method’s ability to perform well despite reduced existing knowledge about the causal gene and disease.

**Figure 4:**
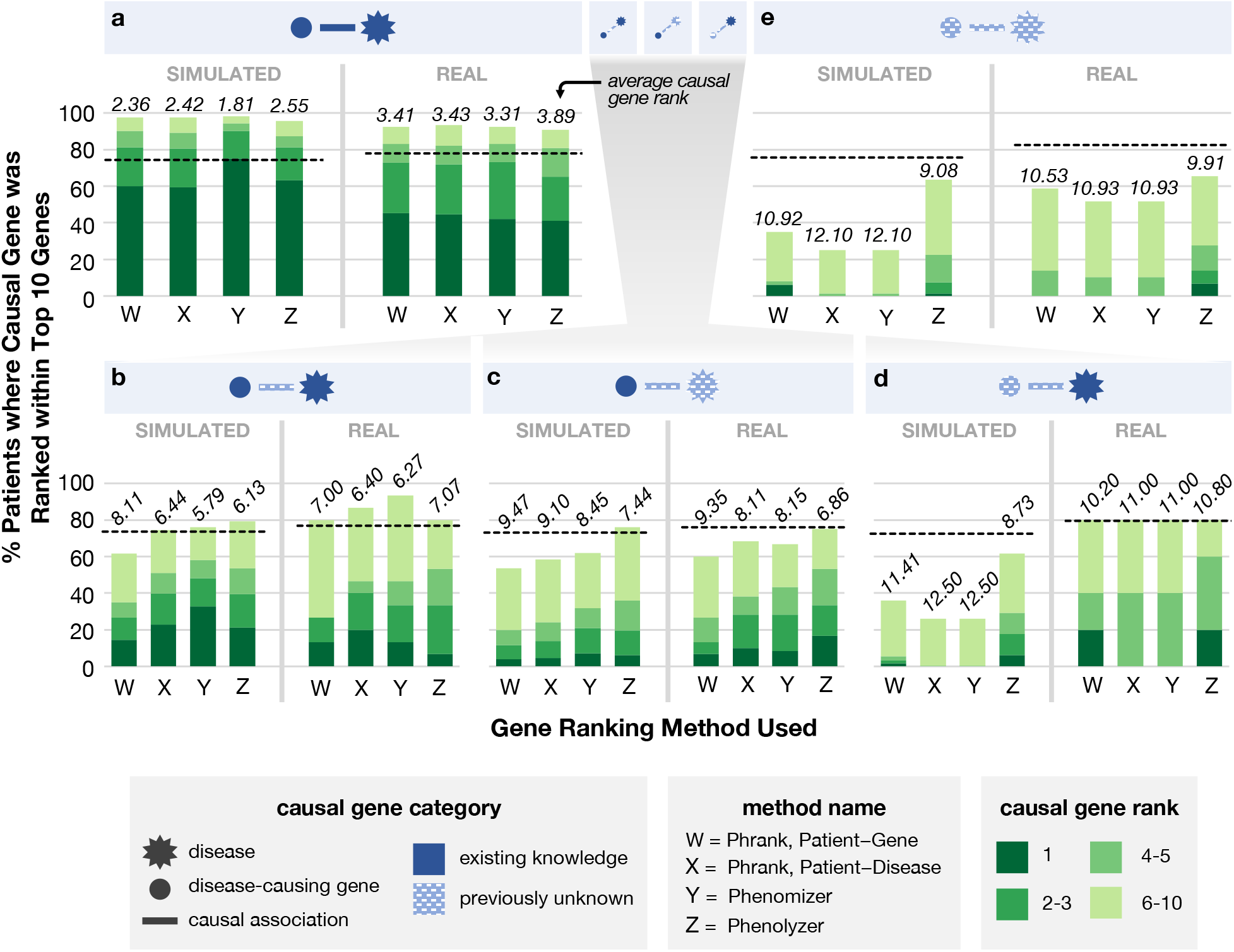
Ability of computational approaches to rank causal genes differs across disease–gene categories. We group simulated patients and real-world UDN patients into five categories based on their type of causal gene–disease association (patient counts in Table 1). These categories, described in detail in Figure 1b, are illustrated in the blue header bars above each plot and ordered decreasingly from left to right by the amount of existing knowledge of the association in the underlying knowledge graph. We run four gene ranking methods on the positive phenotype terms and the candidate gene list for each simulated and real-world patient within each causal gene–disease category, and we show here the ability of these methods to correctly rank each patient’s causal gene within the top 10 ranked genes. The average rank of the causal gene is italicized above each bar. Dashed lines denote the average percent of patients where the causal gene appeared in the top 10 across ten random rankings of the candidate genes.

### Novel syndromes and disease genes represent greatest challenge

All four gene prioritization tools perform well when the patient’s causal gene-disease relationship is in the knowledge graph (Figure 4a). Specifically, all methods rank the causal gene in the top 3 in approximately 75% of simulated and real-world patients, with the causal gene ranked first in over 60% of simulated and over 40% of real-world patients by all methods. However, all methods perform incrementally worse as less information about the simulated patient’s causal gene-disease relationship is present in the knowledge graph. In patients with a known disease caused by a gene previously unassociated with that disease (Figure 4b), the average rank of the causal gene is 5.79 at best. When the patient has a novel disease, even when the causal gene has some other existing disease associations (Figure 4c), performance further declines; the average rank of the causal gene is only 6.86 for the highest performing method. The most difficult scenarios occur when the patient’s causal gene has never been associated with any disease. When the patient’s disease is known (Figure 4d), the average rank of the causal gene is 8.73 for the highest performing method, and when both the disease and causal gene are unknown (Figure 4e), the average rank is 9.08 at best.

Phrank and Phenomizer, which both exclusively use phenotype–phenotype, phenotype–gene and phenotype–disease annotations, excel in settings where the patient’s causal gene and disease are in the knowledge graph (Figure 4a,b). Phenolyzer, which leverages gene–gene associations in addition, outperforms the other two algorithms when either the causal gene or disease are unknown (Figure 4c,d). Indeed, when both the causal gene and disease are unknown (Figure 4e), Phenolyzer ranks the causal gene 9.08 for simulated patients on average whereas both versions of Phrank and Phenomizer rank the causal gene significantly lower, at 10.92, 12.10, and 12.10 respectively on average (all *P* values *<* 2.89 *\** 10^*−*17^). This suggests that the consideration of gene–gene associations may help gene prioritization algorithms generalize to settings where the causal gene has not been previously associated with a disease. When algorithms are evaluated on all patients together rather than separately by novelty category, the general performance decline for all methods across categories as well as relative differences in performance within each novelty category are obfuscated (Supplemental Figure 1).

### Simulation pipeline components are key to simulating realistic, challenging patients

To determine the importance of each component in our simulation pipeline (Figure 2), we run our pipeline using only subsets of these components and then evaluate how well Phrank, the fastest of the three gene prioritization algorithms we evaluated, is able to rank causal genes in the resultant simulated patients. In all ablations, we set the probability of sampling each candidate gene module to be uniform. As expected, we find that the gene prioritization task is easiest when all of the phenotype-altering components and distractor gene modules (as illustrated in Figure 2) are excluded from our simulation pipeline, that is, when candidate genes are selected randomly and phenotype terms do not undergo corruption, dropout, or augmentation with phenotypic noise (Figure 5a). In this setting, the diagnostic gene appears at rank 1.855 on average. The task becomes significantly more difficult when only phenotype-altering components are added (average causal gene rank drops to 1.955; *P* value *<* 0.001) and even more difficult when only distractor gene modules are added (average causal gene rank of 2.570; *P* value *<* 0.001). The task is most difficult when the complete pipeline is used (average causal gene rank of 2.936; *P* value *<* 0.001), suggesting that both the phenotype- and gene-based components of our simulation pipeline contribute to the generation of realistic, challenging rare disease patients.

**Figure 5:**
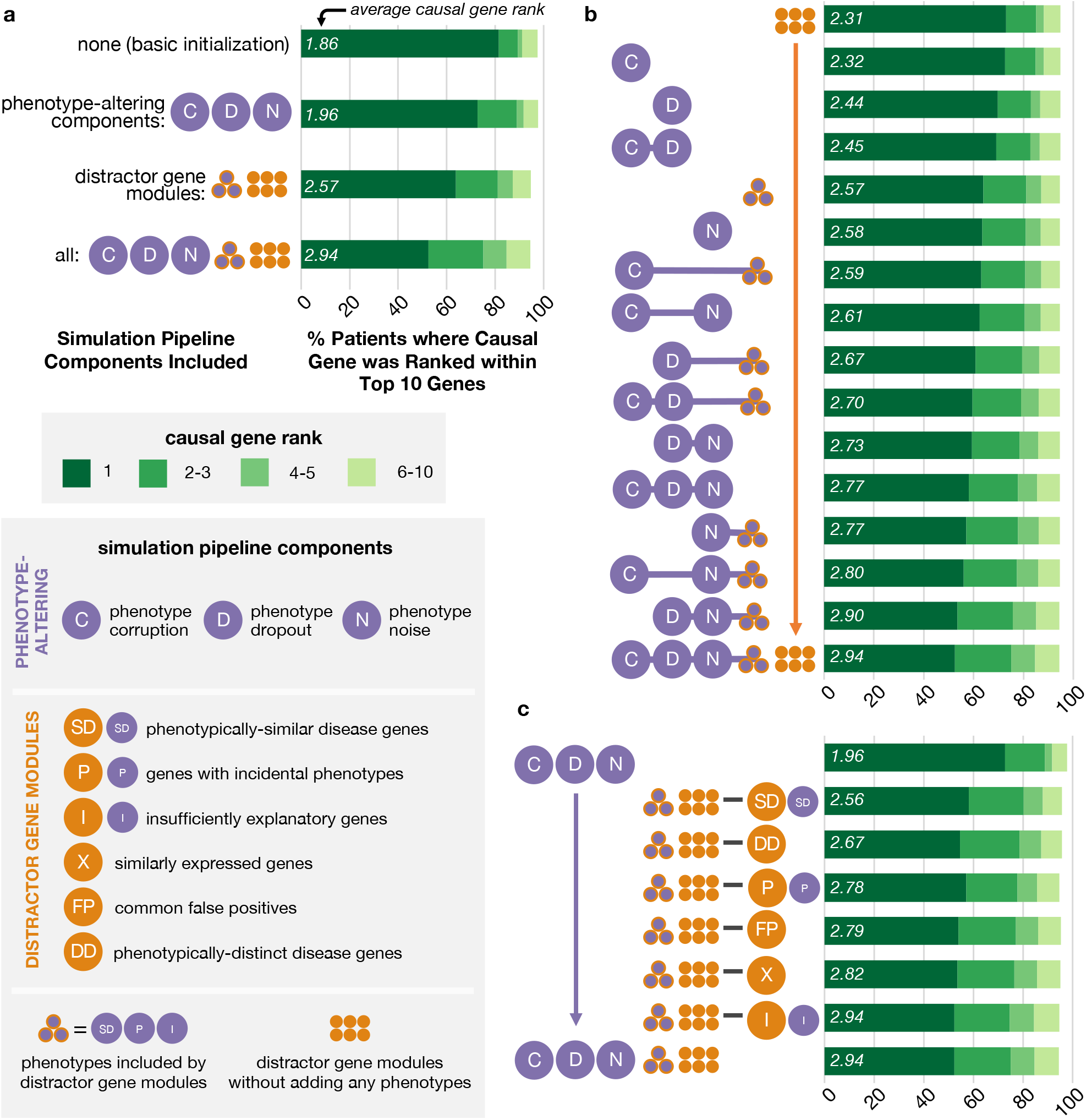
Pipeline components increase the difficulty of causal gene identification in simulated patients. We run a gene prioritization algorithm on patients simulated by our pipeline when varying subsets of pipeline components are included. We report the fraction of simulated patients where the causal gene was prioritized within the top 10 ranked genes (horizontal axis for all plots) when different components of the simulation pipeline are included (vertical axis for all plots). The average rank of the causal gene is listed in italics at the base of each bar. We show gene prioritization performance on simulated patients produced when the following components are included in the simulation pipeline: **a**. no phenotype-nor gene-based components (i.e., candidate genes sampled randomly and phenotype terms unaltered from initialization), all standalone phenotype-altering components alone, all distractor gene modules alone, or all pipeline components together; **b**. a “gene-only” version of distractor gene modules and each possible combination of subsets of phenotype-altering components; **c**. all three standalone phenotype-altering components and all but one distractor gene module at a time. Note that in b, horizontal lines in the vertical axis labels are for visual clarity, whereas in c, horizontal lines in the vertical axis signify set difference.

We next measure how each phenotype-altering component—phenotype corruption, dropout, noise, and phenotypes added by the distractor gene modules—impacts the difficulty of the gene prioritization task (Figure 5b). To this end, we include a “gene-only” version of all distractor gene modules in the simulation pipeline, and we vary whether the distractor gene modules add associated phenotypes and whether phenotype terms undergo corruption, dropout, or noise augmentation. When we restrict to using only one phenotype-altering component at a time, we find that the component that increases the difficulty of the gene prioritization task the most is phenotype noise (average causal gene rank 2.583), followed by phenotypes added by distractor gene modules, phenotype dropout, and phenotype corruption (average causal gene ranks 2.570, 2.435, and 2.316 respectively). Including either the distractor gene-associated phenotypes or phenotype noise alone increases the difficulty of the gene prioritization task more than including both phenotype dropout and corruption together (average causal gene rank 2.448). However, we confirm that including these latter two phenotype-altering components in addition to either one or both of the distractor gene phenotypes and phenotype noise does in fact increase the difficulty of the task more than if they were excluded. In general, adding additional phenotype-altering components makes the gene prioritization task progressively more difficult, and as expected, the most difficult combination is when all phenotype components are included. As before, we find that when the two strongest phenotype-altering components (noisy and distractor gene-associated phenotypes) are applied together, the gene prioritization task is more difficult than when certain sets of three phenotype-altering components are used (average causal gene rank of 2.773 versus average causal gene ranks of 2.767 and 2.702).

Finally, we perform an ablation of each of the six distractor gene modules by removing a single module at a time (Figure 5c). When each distractor gene module is removed, the number of genes that would have been sampled from that module are instead sampled randomly to ensure that the total number of candidate genes for each patient is constant. Removing the module that generates phenotypically-similar disease genes or the module that generates phenotypically-distinct disease genes make the gene prioritization task substantially easier for Phrank relative to the individual exclusion of other distractor gene modules (average causal gene rank of 2.564 and 2.666 respectively). This is intuitive because Phrank’s gene prioritization approach is based on disease–phenotype and gene–phenotype associations. We suspect that the other modules in our pipeline may generate distractor genes that are more challenging for gene prioritization algorithms that explicitly leverage cohort-based mutational recurrence, gene expression, and other additional data. Nevertheless, we confirm that removal of every gene module except for the insufficiently explanatory gene module makes the gene prioritization task easier. There are fewer insufficiently explanatory candidate genes in the ablation patient cohort (Supplemental Figure 2), which may explain why their removal does not change gene prioritization performance. Furthermore, removal of all distractor gene modules together makes the task much easier compared to the removal of any individual module, demonstrating that no one module is solely responsible for generating the distractor genes that increase the difficulty of causal gene prioritization in simulated rare disease patients.

## Discussion

In this work, we developed a flexible framework for simulating difficult-to-diagnose patients with genetic disorders like those profiled in the Undiagnosed Diseases Network (UDN). Key features of our framework include: (i) jointly modeling patients’ genotype and phenotype, (ii) capturing imprecision in real-world clinical workups by corrupting, excluding, and adding noise to patients’ recorded symptoms, and (iii) simulating patients with novel causal gene–disease associations relative to an established knowledge graph, to emulate the challenging task of diagnosing a previously unpublished disorder. Our framework can generate a phenotypically diverse cohort that is representative of all rare diseases characterized in Orphanet, and these simulated patients can be freely shared without privacy concerns.

Our simulated patients are represented as sets of phenotypes and candidate genes, rather than candidate variants. Variant-level properties, such as variant inheritance patterns, functional impacts, and cohort-based frequencies, are considered only indirectly, as we assume that a patient’s candidate genes have been clinically shortlisted because compelling variants have implicated or lie within them [9, 20]. Some variants, such as regulatory variants or larger indels, may impact multiple genes simultaneously. In addition, real-world patients may have two or more genes contributing to their presenting disorder(s). Although our current framework generates patients with monogenic conditions, it could be extended to simulate patients with more than one causal gene; this will become more feasible at scale as the number of rare, multigenic diseases curated in Orphanet increases. As the biomedical data leveraged by prioritization methods diversifies, our framework could be extended to reflect these additions, for instance, by incorporating distractor genes that are highly constrained across human populations and/or phenotypic noise inspired by errors in machine learning-based parsing of clinical notes. Similarly, by building upon the published literature, our work is subject to biases inherent in the field of clinical genetics research writ large, including the historical overrepresentation of individuals of European descent and the diseases that affect them; indeed, we expect that the dataset and methods we present in this paper could be used to further interrogate these biases, for example by examining the differential performance of gene-prioritization tools on diseases that more often affect underrepresented populations.

Finding and annotating “novel” gene–disease associations—defined in our framework as those published post-2015—required significant manual review of public databases and literature. Databases that curate these associations (e.g., Orphanet, HPO) are often missing publication or discovery dates and are not updated in real time, and so an indeterminate lag exists between causal gene–disease associations being present in the literature and being reflected in these knowledge bases. To fairly evaluate gene prioritization methods across the disease novelty categories we describe in Figure 1b, we ensured that all tested methods accessed solely our time stamped knowledge graph from 2015; only methods that used or could be reimplemented to exclusively use this form of input data were included. Indeed, due to their reliance on statically curated data, these methods may have misprioritized real-world patients’ gene–disease associations that were “known” by 2015 but had yet to be incorporated into the knowledge graph, reflecting an expected and ongoing hindrance for diagnosing present-day patients (Figure 4a). We suspect that diagnostic tools that frequently mine the literature for new gene–phenotype–disease associations would excel at diagnosing patients with known causal genes and known diseases relative to tools that rely on structured rare disease databases [21, 22].

We found that the algorithm that was most effective at finding novel disease-causing genes performed relatively poorly at diagnosing patients with known causal genes and diseases, underscoring the importance of evaluating performance separately across distinct novelty categories. Given these findings, clinicians may opt to use certain computational tools earlier in the diagnostic process and move to research-oriented tools only in cases where a novel disease-causing gene is suspected.

We expect that the simulated patients produced via our framework can be leveraged for a wide range of applications [23]. As we demonstrate here, the simulated patients can enable a uniform evaluation of existing gene prioritization tools on a representative patient cohort. Developers can also internally validate and improve their tools by separately evaluating them on simulated patients across novelty categories and distractor gene categories. Another application area for our pipeline will be the generation of training data for machine learning algorithms. As the promise of machine learning solutions in the clinic grows, access to large-scale datasets of relevant clinical data will be essential [24]. We suspect that simulated patients such as those yielded by our method may provide invaluable training data for machine learning models for rare disease diagnosis, which would expose algorithms to data from diverse genetic disorders while reflecting realistic clinical processes.

## Data Availability

The simulated patient dataset as well as all intermediate data used in its creation are shared with the research community via Harvard Dataverse at the DOI: https://doi.org/10.7910/DVN/ANFOR3. Anonymized UDN data has been deposited in dbGaP (accession phs001232) and PhenomeCentral. Phenotypes and causal variants and genes related to UDN diagnoses are also shared publicly in ClinVar: ncbi.nlm.nih.gov/clinvar/submitters/505999/.

https://github.com/EmilyAlsentzer/rare-disease-simulation

## Code availability

The code to reproduce results, together with documentation and examples of usage, can be found at https://github.com/EmilyAlsentzer/rare-disease-simulation.

## Acknowledgements

E.A. is supported by a Microsoft Research PhD Fellowship. S.F. is supported by award Number T32GM007753 from the National Institute of General Medical Sciences. M.L. is supported by T32HG002295 from the National Human Genome Research Institute and a National Science Foundation Graduate Research Fellowship.

UDN research reported in this manuscript was supported by the NIH Common Fund, through the Office of Strategic Coordination/Office of the NIH Director under Award Number(s) U01HG007709, U01HG010219, U01HG010230, U01HG010217, U01HG010233, U01HG010215, U01HG007672, U01HG007690, U01HG007708, U01HG007703, U01HG007674, U01HG007530, U01HG007942, U01HG007943, U01TR001395, U01TR002471, U54NS108251, and U54NS093793. The content is solely the responsibility of the authors and does not necessarily represent the official views of the National Institutes of Health.

## Authors contribution

E.A., S.F, S.K., and I.K. designed the study. E.A. and S.F. implemented the simulation pipeline and gene prioritization algorithms, and E.A. conducted the validation of simulated patients and ablation analysis. M.L. assisted with the UMAP analysis. E.A., S.F., and S.K. wrote the manuscript.

## Competing interests

The authors declare no competing interests.

### Undiagnosed Diseases Network Consortium

Undiagnosed Diseases Network: Maria T. Acosta, Margaret Adam, David R. Adams, Justin Alvey, Laura Amendola, Ashley Andrews, Euan A. Ashley, Mahshid S. Azamian, Carlos A. Bacino, Guney Bademci, Ashok Balasubramanyam, Dustin Baldridge, Jim Bale, Michael Bamshad, Deborah Barbouth, Pinar Bayrak-Toydemir, Anita Beck, Alan H. Beggs, Edward Behrens, Gill Bejerano, Hugo J. Bellen, Jimmy Bennet, Beverly Berg-Rood, Jonathan A. Bernstein, Gerard T. Berry, Anna Bican, Stephanie Bivona, Elizabeth Blue, John Bohnsack, Devon Bonner, Lorenzo Botto, Brenna Boyd, Lauren C. Briere, Elly Brokamp, Gabrielle Brown, Elizabeth A. Burke, Lindsay C. Burrage, Manish J. Butte, Peter Byers, William E. Byrd, John Carey, Olveen Carrasquillo, Thomas Cassini, Ta Chen Peter Chang, Sirisak Chanprasert, Hsiao-Tuan Chao, Gary D. Clark, Terra R. Coakley, Laurel A. Cobban, Joy D. Cogan, Matthew Coggins, F. Sessions Cole, Heather A. Colley, Cynthia M. Cooper, Heidi Cope, William J. Craigen, Andrew B. Crouse, Michael Cunningham, Precilla D’Souza, Hongzheng Dai, Surendra Dasari, Joie Davis, Jyoti G. Dayal, Matthew Deardorff, Esteban C. Dell’Angelica, Katrina Dipple, Daniel Doherty, Naghmeh Dorrani, Argenia L. Doss, Emilie D. Douine, Laura Duncan, Dawn Earl, David J. Eckstein, Lisa T. Emrick, Christine M. Eng, Cecilia Esteves, Marni Falk, Liliana Fernandez, Elizabeth L. Fieg, Paul G. Fisher, Brent L. Fogel, Irman Forghani, William A. Gahl, Ian Glass, Bernadette Gochuico, Rena A. Godfrey, Katie Golden-Grant, Madison P. Goldrich, Alana Grajewski, Irma Gutierrez, Don Hadley, Sihoun Hahn, Rizwan Hamid, Kelly Hassey, Nichole Hayes, Frances High, Anne Hing, Fuki M. Hisama, Ingrid A. Holm, Jason Hom, Martha Horike-Pyne, Alden Huang, Yong Huang, Wendy Introne, Rosario Isasi, Kosuke Izumi, Fariha Jamal, Gail P. Jarvik, Jeffrey Jarvik, Suman Jayadev, Orpa Jean-Marie, Vaidehi Jobanputra, Lefkothea Karaviti, Jennifer Kennedy, Shamika Ketkar, Dana Kiley, Gonench Kilich, Shilpa N. Kobren, Isaac S. Kohane, Jennefer N. Kohler, Deborah Krakow, Donna M. Krasnewich, Elijah Kravets, Susan Korrick, Mary Koziura, Seema R. Lalani, Byron Lam, Christina Lam, Grace L. LaMoure, Brendan C. Lanpher, Ian R. Lanza, Kimberly LeBlanc, Brendan H. Lee, Roy Levitt, Richard A. Lewis, Pengfei Liu, Xue Zhong Liu, Nicola Longo, Sandra K. Loo, Joseph Loscalzo, Richard L. Maas, Ellen F. Macnamara, Calum A. MacRae, Valerie V. Maduro, Rachel Mahoney, Bryan C. Mak, May Christine V. Malicdan, Laura A. Mamounas, Teri A. Manolio, Rong Mao, Kenneth Maravilla, Ronit Marom, Gabor Marth, Beth A. Martin, Martin G. Martin, Julian A. Martínez-Agosto, Shruti Marwaha, Jacob McCauley, Allyn McConkie-Rosell, Alexa T. McCray, Elisabeth McGee, Heather Mefford, J. Lawrence Merritt, Matthew Might, Ghayda Mirzaa, Eva Morava, Paolo M. Moretti, Mariko Nakano-Okuno, Stan F. Nelson, John H. Newman, Sarah K. Nicholas, Deborah Nickerson, Shirley Nieves-Rodriguez, Donna Novacic, Devin Oglesbee, James P. Orengo, Laura Pace, Stephen Pak, J. Carl Pallais, Christina GS. Palmer, Jeanette C. Papp, Neil H. Parker, John A. Phillips III, Jennifer E. Posey, Lorraine Potocki, Barbara N. Pusey, Aaron Quinlan, Wendy Raskind, Archana N. Raja, Deepak A. Rao, Anna Raper, Genecee Renteria, Chloe M. Reuter, Lynette Rives, Amy K. Robertson, Lance H. Rodan, Jill A. Rosenfeld, Natalie Rosenwasser, Francis Rossignol, Maura Ruzhnikov, Ralph Sacco, Jacinda B. Sampson, Mario Saporta, Judy Schaechter, Timothy Schedl, Kelly Schoch, C. Ron Scott, Daryl A. Scott, Vandana Shashi, Jimann Shin, Edwin K. Silverman, Janet S. Sinsheimer, Kathy Sisco, Edward C. Smith, Kevin S. Smith, Emily Solem, Lilianna Solnica-Krezel, Ben Solomon, Rebecca C. Spillmann, Joan M. Stoler, Jennifer A. Sullivan, Kathleen Sullivan, Angela Sun, Shirley Sutton, David A. Sweetser, Virginia Sybert, Holly K. Tabor, Amelia L. M. Tan, Queenie K.-G. Tan, Mustafa Tekin, Fred Telischi, Willa Thorson, Cynthia J. Tifft, Camilo Toro, Alyssa A. Tran, Brianna M. Tucker, Tiina K. Urv, Adeline Vanderver, Matt Velinder, Dave Viskochil, Tiphanie P. Vogel, Colleen E. Wahl, Melissa Walker, Stephanie Wallace, Nicole M. Walley, Jennifer Wambach, Jijun Wan, Lee-kai Wang, Michael F. Wangler, Patricia A. Ward, Daniel Wegner, Monika Weisz-Hubshman, Mark Wener, Tara Wenger, Katherine Wesseling Perry, Monte Westerfield, Matthew T. Wheeler, Jordan Whitlock, Lynne A. Wolfe, Kim Worley, Changrui Xiao, Shinya Yamamoto, John Yang, Diane B. Zastrow, Zhe Zhang, Chunli Zhao, Stephan Zuchner

## Methods

### 1 Simulated Patient Initialization

We simulate patients for each of the 2,134 diseases in Orphanet [19] (orphadata.org, accessed October 29, 2019) that do not correspond to a group of clinically heterogeneous disorders (i.e., “Category” classification), have at least one associated phenotype, and have at least one causal gene. For Orphanet diseases that were missing either a causal gene or phenotypes (but not both), and were listed as being a ‘clinical subtype’, ‘etiological subtype’, or ‘histopathological subtype’ of another Orphanet disease that did have a causal gene and/or phenotypes, we imported the causal gene and/or phenotypes from the parent disease. For each patient, the gene set is initialized with the known causal disease gene (mapped to its Ensembl identifier); the age is randomly sampled from the age ranges associated with the disease (e.g., “infant”); and positive and negative phenotype terms from the Human Phenotype Ontology [25] (HPO, version 2019) are added with probabilities *Pr*(*term*|*disease*) and 1–*Pr*(*term*|*disease*) respectively, where *Pr*(*term*|*disease*) is provided in Orphanet and corresponds to the observed prevalence of a specific phenotype term presenting in patients with the disease.

### 2 Modeling Diagnostic Process Imprecision

To mimic real-world patients’ partially observed phenotypes, we perform phenotype dropout where each positive and negative phenotype term is removed from the simulated patient with probabilities Pr(positive dropout) and Pr(negative dropout), respectively set to 0.7 and 0.2 in our implementation. We also perform phenotype corruption to replace specific phenotype terms (e.g., “arachnodactyly”) with their less precise parent terms (e.g., “long fingers”) in the HPO ontology. Positive and negative phenotypes annotated to each patient are corrupted with probabilities Pr(positive corruption) and Pr(negative corruption), each set to 0.15 in our implementation. Finally, to model unrelated symptoms and comorbidities that would be present in real-world patients, we introduce phenotype noise by sampling new HPO phenotype terms that were mapped from ICD-10 billing codes from a large medical insurance claims database using the Unified Medical Language System (UMLS) crosswalk [26]. Positive phenotype terms are sampled with probabilities proportional to their prevalence in corresponding age-stratified populations (i.e., infants are defined as 0-1 years, children are 2-11 years, adolescents are 12-18 years, adults are 19-64 years, and seniors are 65+ years), and negative phenotype terms are sampled from the same corresponding age-stratified populations at random.

### 3 Distractor Gene Modules

In order to mimic the typical diagnostic process where numerous potentially disease-causal genes must be manually reviewed by a clinical team, we generate a set of 1 + *N*_*G*_ highly plausible, yet ultimately non-causal, genes for each patient, where *N*_*G*_ is drawn from a Poisson distribution parameterized by *λ*. In our implementation, the tunable parameter *λ* is set to the mean number of candidate genes considered in real-world patients with undiagnosed genetic conditions (see Methods “Preprocessing Real Patient Data” below). These *N*_*G*_ genes are generated from the following six distractor gene modules with probabilities 0.33, 0.42, 0.05, 0.09, 0.08, and 0.03 respectively, which we set in our implementation based on the approximate frequency of each distractor gene type in real-world patients with undiagnosed diseases; these parameters can be customized by the user. Each distractor gene module contributes one gene to the simulated patient’s candidate gene list, and three distractor gene modules simultaneously add phenotype terms related to the added gene.

#### 1. Phenotypically-similar disease genes

First, we identify genes causing “distractor” Mendelian diseases in Orphanet that have overlapping phenotype terms with the patient’s true disease (Figure 2b, dark green). We categorize the phenotype terms associated with the distractor disease as “obligate”, “strong”, “weak”, or “excluded” if their prevalence in patients with that disease is 100%, 80-99%, 1-29% or 0% respectively. We require that all distractor diseases have at least one obligate phenotype term and/or at least one excluded phenotype term, or that all phenotype terms that overlap with the true disease of interest are weak. We add the causal gene for the distractor disease to the simulated patient’s set of candidate genes and add phenotype terms that overlap between the distractor and true disease to the simulated patient’s positive phenotype set. To ensure that these genes are challenging distractors but definitively non-causal, we add some excluded phenotypes to the simulated patient’s set of positive phenotypes and some obligate phenotypes to the simulated patient’s set of negative phenotypes. For distractor diseases with no associated obligate or excluded phenotypes and only weak overlapping phenotypes with the true disease, we instead add some strong, non-overlapping phenotypes to the simulated patient’s negative phenotypes. At each of these steps, 1 + *N*_*P*_ phenotype terms are added to the simulated patient’s positive or negative phenotype sets, where *N*_*P*_ is drawn from a Poisson distribution parameterized by *λ*. In our implementation, we set *λ* such that simulated patients and real-world patients with undiagnosed diseases have approximately the same number of annotated phenotype terms on average (see Figure 3 and Methods “Preprocessing Real Patient Data” below).

#### 2. Phenotypically-distinct disease genes

Since any variants in known disease genes tend to be investigated during the diagnostic process, we also add genes causing Mendelian diseases that do not have any phenotypic overlap with the patient’s true disease (Figure 2b, yellow) [27].

#### 3. Insufficiently explanatory genes

Genes that are not yet known to be disease-causing but are associated with a subset of the patient’s disease-relevant phenotypes are also strong candidates for further diagnostic investigation. To generate such insufficiently explanatory genes, we first curate a set of non-disease genes as the set of all genes from DisGeNET [28] (accessed April 16, 2019, https://www.disgenet.org/downloads/all gene disease associations.tsv) and excluding any genes causally associated with a disease in Orphanet or in HPO Annotation [29, 30] (accessed February 12, 2019, http://compbio.charite.de/jenkins/job/hpo.annotations.monthly/ALL_SOU_RCES_ALL_FREQUENCIES diseases to genes to phenotypes.txt). We add non-disease genes that are associated with a strict subset of low prevalence phenotypes from the simulated patient’s true disease (Figure 2b, light orange). We then add 1 + *N*_*P*_ of the gene’s phenotype terms to the simulated patient’s positive phenotype set if none are already present, with *N*_*P*_ defined as above.

#### 4. Genes associated with incidental phenotypes

Naturally occurring phenotypic variance present across healthy individuals can be incorrectly considered to be relevant to a patient’s disease during diagnostic evaluations. To include genes causing these nonsyndromic phenotypes, we add non-disease genes associated only with phenotypes that do not overlap with the simulated patient’s true disease (Figure 2b, purple). We add some of the gene’s phenotypes to the simulated patient’s positive phenotype set as before.

#### 5. Similarly expressed genes

We also add genes with similar tissue expression as the patient’s causal disease gene (Figure 2b, dark orange), as a candidate gene’s expression in relevant tissues is considered as supporting experimental evidence for a gene-disease association in clinical evaluations [31]. For each gene, we compute its average tissue expression in transcripts per million in each of 54 tissue types profiled in GTEx [32] (accessed October 29, 2019, https://gtexportal.org/home/datasets/GTEx_Analysis_2017-06-05_v8_RNASeQCv1.1.9_gene_median_tpm.tsv). For each tissue type, we linearly 0,1-normalize the per-gene expression values such that the gene with the lowest expression in that tissue type is assigned a value of 0 and the gene with the highest expression in that tissue type is assigned a value of 1. We compare each gene’s normalized tissue expression vector to the simulated patient’s causal gene’s tissue expression vector using cosine similarity. We select one of the top 100 most similar genes with probability proportional to its tissue expression similarity, excluding known disease genes with phenotypic overlap with the simulated patient’s true disease.

#### 6. Common false positive genes

Finally, we add genes from the FrequentLy mutAted GeneS (FLAGS) database [33] with probabilities proportional to the number of rare functional variants affecting these genes in general populations, as computational pipelines tend to frequently prioritize these genes due to their length and variational excess.

### 4 Preprocessing Real Patient Data

We selected all patients from the Undiagnosed Diseases Network (UDN) with a molecular diagnosis as of March 19, 2020. Each patient is annotated with a set of positive and negative HPO phenotype terms and a set of strong candidate genes that were considered by clinical teams who handled each case. For each of the 362 diagnosed patients who received genomic sequencing through the UDN, we augment their candidate gene lists with disease-associated and other clinically-relevant genes listed on their clinical sequencing reports [27]. Where possible, patients’ gene lists were further augmented with genes prioritized by the Brigham Genomic Medicine pipeline [34]. We map all genes to Ensembl identifiers, discard prenatal phenotype terms related to the mother’s pregnancy, and exclude patients with fewer than five candidate genes. The final cohort includes 248 patients.

The Undiagnosed Diseases Network study is approved by the National Institutes of Health institutional review board (IRB), which serves as the central IRB for the study (Protocol 15HG0130). All patients accepted to the UDN provide written informed consent to share their data across the UDN as part of a network-wide informed consent process.

### 5 Comparing Simulated Patients to Real Patients

Of the 248 diagnosed UDN patients that we consider, only 121 patients were diagnosed with a disease in Orphanet that we were able to model (see Section 1 above). We construct a disease-matched cohort of 2,420 simulated patients by selecting, for each of these 121 UDN patients, 20 simulated patients with the same disease. We first visualize positive phenotype term similarities between real and simulated patients using non-linear dimension reduction via a Uniform Manifold Approximation and Projection (UMAP) plot [35]. We also compare each real patient to all simulated patients in the disease-matched cohort by computing pairwise Jaccard similarities ranging from 0 to 1 inclusively between the positive phenotype terms annotated to each real patient and the positive phenotype terms annotated to each simulated patient. For each real patient, we then rank all simulated patients from highest to lowest Jaccard similarity and analyze those simulated patients’ corresponding diseases to assess whether simulated patients are as phenotypically specific to each disease as real patients are. Finally, we evaluate the average Jaccard similarity between the query real-world patient and the top 10 retrieved simulated patients with the same disease compared to the average Jaccard similarity between the query and the top 10 real-world patients with a different disease using a one-sided Wilcoxon signed-rank test implemented in the SciPy Stats library. We perform the Shapiro-Wilk test from the SciPy Stats library to test for normality.

### 6 Evaluating Gene Prioritization Tools on Novel Diseases

To model novel genetic conditions in our simulated patients, we leverage a knowledge-graph (KG) of gene–gene, gene–disease, gene–phenotype, and phenotype–disease annotations from Phenolyzer [11] that is time-stamped to February 2015 (obtained from github.com/WGLab/phenoly zer/tree/ecec7410729276859b9023a00f20e75c2ce58862) and the HPO-A ontology [30] time stamped to January 2015 (obtained from github.com/drseb/HPO-archive/tree/master/hpo.annot ations.monthly/2015-01-28 14-15-03/archive/annotation and github.com/drseb/HPO-archive/tree/master/2014-2015/2015 week 4/annotations/artefacts). Phenotype-phenotype annotations are from the 2019 HPO ontology. All gene names are mapped to Ensembl IDs, and older phenotype terms are updated to the 2019 HPO ontology. We also manually time-stamp each disease and disease–gene association in Orphanet according to the date of the Pubmed article that reported the discovery; discoveries after February 2015 are considered novel with respect to our KG. Note that the KG time-stamped to February 2015 may not reflect all new information contained in the most recent publications from PubMed, as would be expected for any curated database. We categorize each novel discovery as in Figure 1b. We apply the same process to manually annotate the novelty of disease-gene associations in the real-world UDN cohort.

We reimplement four well-known gene prioritization tools using our 2015 time-stamped KG. We use publicly-available code from https://bitbucket.org/bejerano/phrank and https://github.com/WGLab/phenolyzer to run Phrank [10] and Phenolyzer [11] respectively. We reimplement Phenomizer [12] as described in their paper, as open-source code is not available.

Although the original implementation of Phenomizer randomly samples phenotype terms 100,000 times to generate p-values for each patient–disease similarity score, we use 10,000 random samplings instead, as this was substantially faster, and varying the number of samplings did not impact overall gene rankings. We define a patient–gene match score for Phenomizer as the highest patient–disease match score across all diseases caused by that gene. We report how well each of these tools—both versions of Phrank, Phenolyzer, and Phenomizer—ranked the causal gene for each simulated patient for each different category of novel disorders as outlined in Figure 1b.

## Notes

### Competing Interest Statement

The authors have declared no competing interest.

### Author Declarations

The Undiagnosed Diseases Network study is approved by the National Institutes of Health institutional review board (IRB), which serves as the central IRB for the study (Protocol 15HG0130).

### Summary of Updates

A typo in the abstract was corrected and the formatting for URLs in the paper was updated.

