## Supplementary Information for "Simulation of undiagnosed patients with novel genetic conditions"

### Contents

#### List of Figures

S1 Overall ability of computational approaches to rank causal genes on entire simulated and real-world datasets . . . . . S3
S2 Simulated patients have relatively fewer candidate genes from the Insufficiently Explanatory Gene Module . . . . . S4

### Supplementary Figures

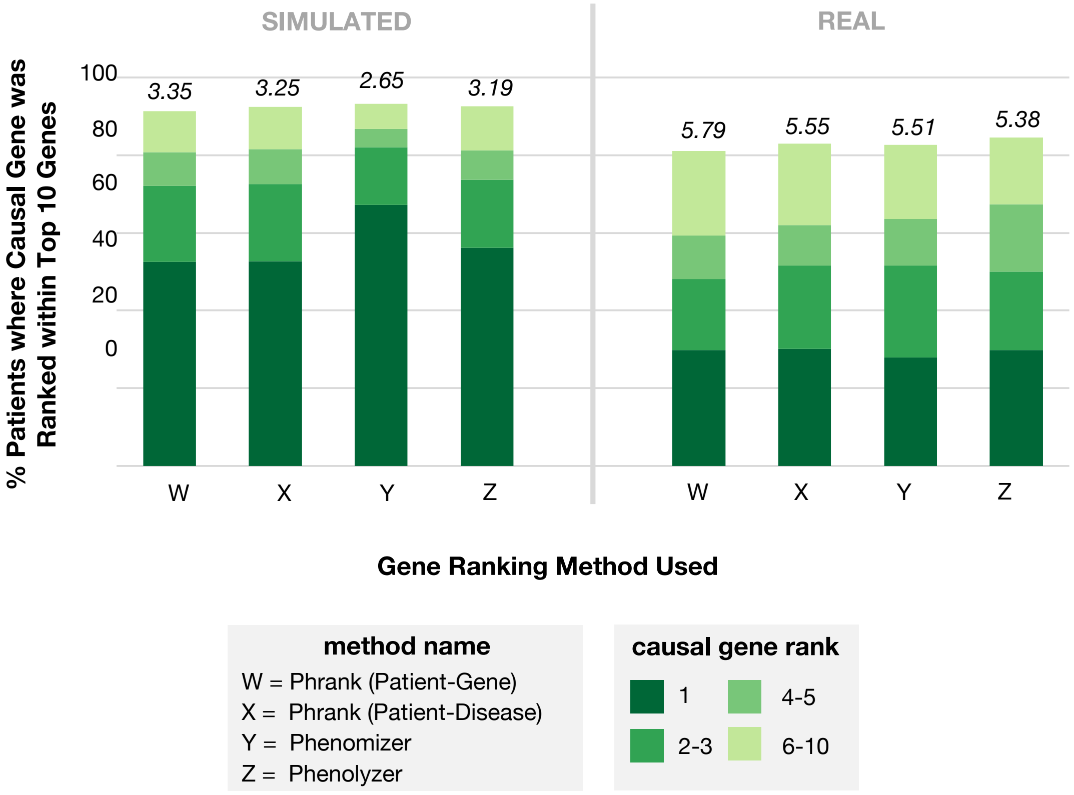


**Figure S1: Overall ability of computational approaches to rank causal genes on entire simulated and real-world datasets.** We run four gene ranking methods on the positive phenotype terms and candidate genes for all simulated and real-world patients in the cohorts, and we show here the ability of these methods to correctly rank each patient's causal gene within the top 10 ranked genes. The average rank of the causal gene is italicized above each bar. Notably, the performance of the algorithms on the entire cohort does not reflect heterogeneity in performance across disease–gene categories as shown in Figure 4.

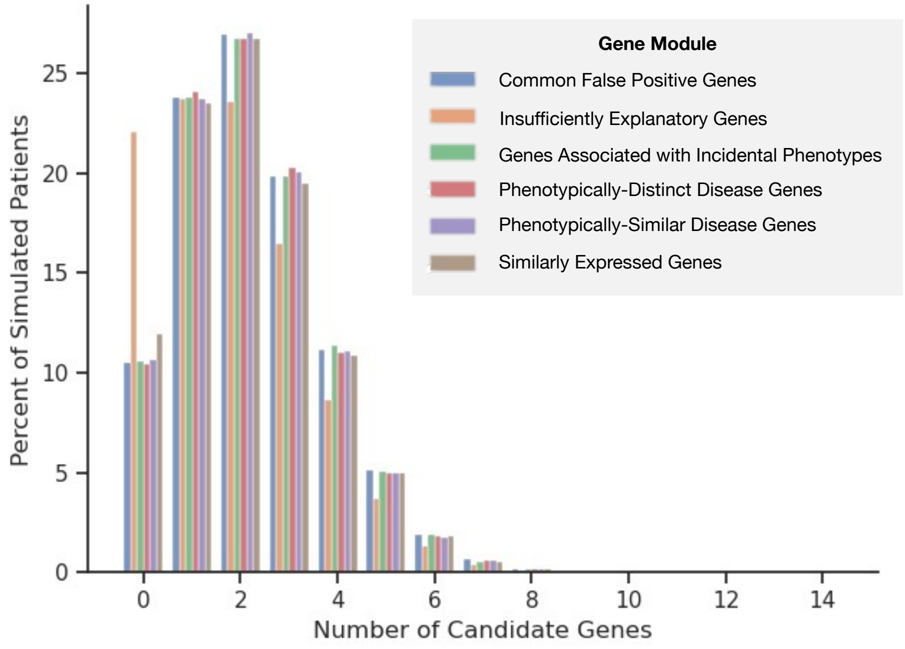


**Figure S2: Simulated patients have relatively fewer candidate genes from the Insufficiently Explanatory Gene Module.** In the gene module ablation experiment, we run the simulation pipeline with an equal probability of sampling each distractor gene module and perform an ablation of each of the six distractor gene modules by removing a single module at a time. Despite sampling each module with equal probability, there are fewer patients with candidate genes added by the Insufficiently Explanatory Gene Module compared to other gene modules in our initial simulated patient cohort. Insufficiently explanatory genes may not be added to a patient if there are no qualifying non-disease genes that are associated with a strict subset of low prevalence phenotypes from the simulated patient's true disease (See Methods). The lower prevalence of Insufficiently Explanatory genes may explain why removal of these genes does not change gene prioritization performance as shown in Figure 5c.
